# An Explainable Advanced Electrocardiography Score for Diastolic Dysfunction - Derivation, Validation and Prognostic Performance

**DOI:** 10.1101/2023.04.19.23288666

**Authors:** Hayden McColl, Abhinayeni Kuhasri, Sabbab Chowdhury, Yalu Wen, Daniel E Loewenstein, Todd T Schlegel, Maren Maanja, Kevin X Yang, Zaidon Al-Falahi, Thomas Lindow, Michelle Price, Elias Fulthorp, Sean Lal, Michele McGrady, Patrick Gladding, Rebecca Kozor, Martin Ugander

## Abstract

**BACKGROUND AND AIMS:** Diastolic dysfunction is a precursor to heart failure with preserved ejection fraction (HFpEF), and early detection by electrocardiography (ECG) would be valuable. We hypothesised that an explainable advanced ECG (A-ECG) score could accurately detect diastolic dysfunction with clinically meaningful diagnostic and prognostic performance.

**METHODS:** A derivation cohort was included after standard 12-lead ECG and echocardiography demonstrating normal systolic function, and either the presence or absence of diastolic dysfunction (≥grade 2, 2025 guidelines). A multivariable machine learning A-ECG diastolic dysfunction score (0-100%, positive score defined as >50%) was derived using elastic net with nested resampling. Performance for identifying diastolic dysfunction was assessed in a separate external validation cohort. Prognostic performance for predicting cardiovascular events including mortality was evaluated in the UK Biobank.

**RESULTS:** A 7-measure A-ECG diastolic dysfunction score derived in the derivation cohort (n=414) performed excellently in the validation cohort (n=3,418, area under the receiver operating characteristics curve [95% confidence interval] 0.87 [0.86-0.88]). Survival analysis using spline modelling adjusted for age, sex, and cardiovascular risk factors in the UK Biobank (n=27,239, 966 events, follow-up 1.9 [0.7–4.4] years, age 66±8 years, 50% female) showed that over the range of the score (0-100%), the hazard ratio for events increased linearly to 4 (p<0.001)

**CONCLUSIONS:** Explainable A-ECG analysis of the standard 12-lead ECG can be used to accurately identify diastolic dysfunction. The score reflecting the continuous probability (0-100%) of having diastolic dysfunction has a linearly increasing and strong adjusted prognostic association with cardiovascular events.

## Introduction

Left ventricular (LV) diastolic dysfunction describes abnormalities in myocardial relaxation, compliance, and filling, and is fundamental to the development of heart failure with preserved ejection fraction (HFpEF)^1, 2^. Diastolic dysfunction may be initially asymptomatic before progressing to symptomatic HFpEF, and may be present for many years before manifesting as overt heart failure^3, 4^. Aggressive management of cardiovascular risk factors including hypertension, diabetes, and obesity has been shown to prevent or reduce disease progression^5^. Due to diastolic dysfunction commonly being asymptomatic before manifesting as HFpEF, earlier identification to better enable risk factor modification is desirable.

Diastolic dysfunction is most commonly assessed using transthoracic echocardiography. However, echocardiographic screening is not feasible on a population scale due to resource-related constraints^6^. As such, a low-cost screening method for diastolic dysfunction would be clinically valuable. Attempts to use the electrocardiogram (ECG) as a screening tool for detecting diastolic dysfunction have shown promise. However, previous studies have relied heavily upon clinical data beyond the ECG, included methods that are either difficult to implement on a largescale due to requirements for dedicated hardware that may not be widely available, or that require the use of artificial intelligence models that can be limited by not being readily explainable^7–9^.

Advanced electrocardiography (A-ECG) is a term describing the application of multivariable statistics to a large number of measures obtained from a combination of digital signal processing analysis methods applied to the standard resting 12-lead ECG^10, 11^. Notably, A-ECG analysis can be performed on the digital ECG raw data recordings stored by any modern ECG vendor. A-ECG analysis of the standard resting 10-sec ECG enables improved diagnostic performance by incorporating not only conventional ECG measurements such as waveform amplitudes, axes and durations, but also measures of derived three-dimensional vectorcardiography (VCG) and waveform complexity via singular value decomposition^11–13^. A-ECG has been able to discriminate between health and disease in several conditions such as coronary artery disease, cardiomyopathy, LV hypertrophy and LV systolic dysfunction, with notably higher diagnostic accuracy compared to conventional ECG interpretation^11, 14, 15^.

However, there have not yet been studies examining the utility of A-ECG in the detection of diastolic dysfunction. A-ECG may have use for screening at-risk asymptomatic patients for diastolic dysfunction in the setting of constrained resources. Such patients might benefit from further investigation and aggressive risk factor management to prevent progression of disease to HFpEF and reduce its globally increasing burden.

Therefore, the aim of this study was to derive and validate an A-ECG score for detecting diastolic dysfunction using the standard resting 12-lead ECG. We hypothesised that combined measures of A-ECG analysis derived from the ECG could be used to accurately identify the presence of diastolic dysfunction from echocardiography, and that this score would have a prognostic association with cardiovascular events in a large outpatient cohort.

## Methods

### Study Design

This was a retrospective cohort study to derive and validate an A-ECG score for the detection of diastolic dysfunction as determined by transthoracic echocardiography (TTE). The derivation cohort consisted of patients who were referred for assessment at an academic outpatient cardiology practice between January, 2018 and March, 2022. External validation was subsequently performed in an independent cohort drawn from a separate healthcare institution. The study received ethics approval from the Sydney Local Health District Ethics Board (X19-0286 & 2019/ETH12064) as well as the Waitemata District Health Board (2023/93783).

Echocardiographic examinations were evaluated and patients classified as having either normal or abnormal diastolic function defined as meeting criteria for either grade 2 or 3 diastolic dysfunction per the 2025 American Society of Echocardiography guidelines. Normal diastolic function was defined as the absence of any required criteria for diastolic dysfunction^16^.

*Derivation cohort*. The derivation cohort inclusion criteria were age >50 years, sinus rhythm, left ventricular ejection fraction (LVEF) ≥55% and a recorded 12-lead ECG within 1 month of echocardiography. Patients were excluded if they had moderate or greater valvular disease, LVEF <55%, hypertrophic cardiomyopathy, previous valvular replacement, coronary artery bypass surgery, or any other cardiac procedures that had required entering the pericardium. Exclusion criteria were ECG abnormalities that might limit A-ECG analysis such as atrial arrhythmias, predominant ventricular ectopic beats or QRS duration >120ms.

*Validation cohort*. The diagnostic validation cohort had a broad inclusion criterion of all patients with either normal diastolic function, or grade 2-3 diastolic dysfunction. Again, patients were excluded for ECG abnormalities which would limit A-ECG analysis. Sensitivity analysis of our score’s performance was then included by the addition of grade 1 diastolic function to our validation cohort. The inclusion flowchart for the validation and derivation cohorts is outlined in the supplementary material.

*Prognosis cohort*. The prognostic performance of the A-ECG score for diastolic dysfunction was evaluated in a cohort from the UK biobank, which is an outpatient population recruited from the general public in the UK. Prognosis was assessed as a composite of time to first cardiovascular event or all-cause mortality following the date of the index ECG. Diagnostic codes (ICD-10) and procedural codes (OPCS-4) classified as events are detailed in the supplementary material. Included subjects had undergone a resting 12-lead ECG as well as a baseline questionnaire detailing their medical, family and psychosocial history. Included subjects were divided into those without pre-existing cardiovascular disease and those with either self-reported cardiovascular disease (stroke, myocardial infarction or angina) or a pre-existing diagnosis of cardiac conditions as detailed by diagnostic and procedural codes (Appendix C). Subjects were excluded if they had ECG abnormalities which would limit analysis, such as significant atrial or ventricular arrhythmia. A subject inclusion flowchart is detailed in the supplementary material.

### Advanced electrocardiography analysis

All ECG recordings were performed by a commercially available ECG system that acquired a standard resting 12-lead ECG of 10 second duration (Mortara Instruments Inc, USA; General Electric, USA). A-ECG analysis and its extensive methods have been detailed previously^11, 17^. In brief, conventional ECG parameters, including all major waveform intervals, axes, and voltages were derived automatically from the ECG recording software. A-ECG parameters were obtained after signal averaging the ECG recording using in-house developed software^11, 13, 17^. Vectorcardiographic measures were obtained from the Frank X, Y, and Z leads derived from the 12-lead ECG using Kors’ regression transformation^18^. QRS– and T-wave complexity measures were quantified from eigenvector lead waveforms calculated using singular value decomposition of the 12-lead ECG^11, 17^.

### Statistical Analysis

Statistical analyses including feature selection procedures were conducted using R version 4.2.0 (R Core Team, R Foundation for Statistical Computing, Vienna, Austria) with key packages tidymodels, glmnet, survival, plotRCS used for data analysis and visualisation ^19–22^. A multivariable A-ECG score for assessing diastolic dysfunction was derived using elastic net logistic regression^23^. Nested resampling was applied to determine cross-validated diagnostic accuracy and obtain 95% confidence intervals (CI). Nested resampling is a method of cross validation and model evaluation that uses two levels of repeated sampling to minimise the risk of overfitting by optimisation bias in a predictive model^24^. The A-ECG score was then used to estimate the probability of a given subject in the study cohort having diastolic dysfunction, with a cut-off predicted probability of >50%. The score calculated the likelihood of having diastolic dysfunction from the elastic net regression as:

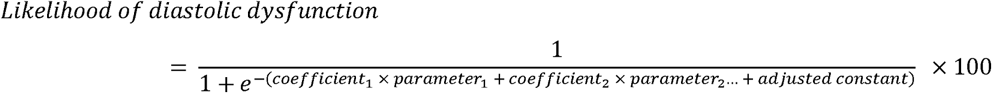

Time-to-event analysis was performed using Kaplan–Meier curves with censoring at the conclusion of follow-up, with subjects grouped into quartiles of their A-ECG score. Restricted cubic spline plots were generated to demonstrate changes in hazard ratio with increasing diastolic dysfunction score as a continuous variable. Multivariable spline analysis was also undertaken to assess the impact of increasing diastolic dysfunction score whilst adjusting for age and other cardiovascular risk factors including hypertension, diabetes mellitus, dyslipidaemia, family history, smoking, and elevated body mass index.

## Results

### Study Population

The derivation cohort consisted of 414 patients, 264 (64%) with normal diastolic function and 150 (36%) with diastolic dysfunction (133 [32%] with grade 2 and 17 [4%] with grade 3 diastolic dysfunction). In the normal diastolic function group, 45% were males and mean age was 62±9 years. In the diastolic dysfunction group, 35% were males and mean age was 77±9 years. Detailed demographic information of the derivation cohort is presented in the supplementary material. The diagnostic validation cohort consisted of 3,418 patients, 2,649 (78%) with normal diastolic function and 769 (22%) with diastolic dysfunction (690 [20%] with grade 2 and 79 [2%] with grade 3 diastolic dysfunction). Detailed demographic information of the validation cohort is presented in Table 1. Sensitivity analysis was performed in an expanded population (total n=4,616) by the addition of patients with grade 1 diastolic dysfunction (n=1,198). Baseline characteristics of this expanded sensitivity analysis cohort are outlined in the supplementary material.

**Table 1.**
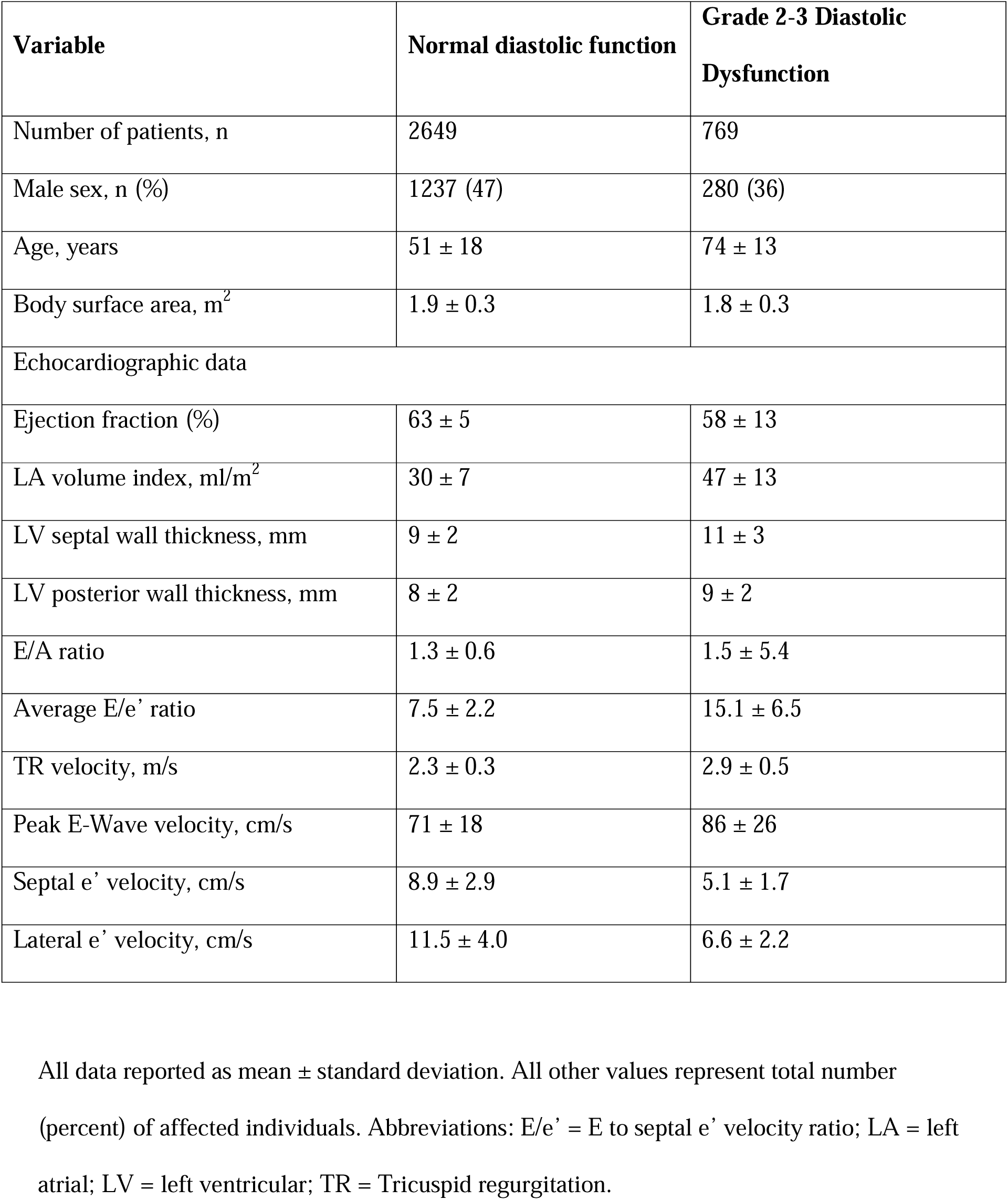
Validation Cohort Patient characteristics.

| Variable | Normal diastolic function | Grade 2-3 Diastolic Dysfunction |
| --- | --- | --- |
| Number of patients, n | 2649 | 769 |
| Male sex, n (%) | 1237 (47) | 280 (36) |
| Age, years | 51 ± 18 | 74 ± 13 |
| Body surface area, m<sup>2</sup> | 1.9 ± 0.3 | 1.8 ± 0.3 |
| Echocardiographic data |  |  |
| Ejection fraction (%) | 63 ± 5 | 58 ± 13 |
| LA volume index, ml/m<sup>2</sup> | 30 ± 7 | 47 ± 13 |
| LV septal wall thickness, mm | 9 ± 2 | 11 ± 3 |
| LV posterior wall thickness, mm | 8 ± 2 | 9 ± 2 |
| E/A ratio | 1.3 ± 0.6 | 1.5 ± 5.4 |
| Average E/e' ratio | 7.5 ± 2.2 | 15.1 ± 6.5 |
| TR velocity, m/s | 2.3 ± 0.3 | 2.9 ± 0.5 |
| Peak E-Wave velocity, cm/s | 71 ± 18 | 86 ± 26 |
| Septal e' velocity, cm/s | 8.9 ± 2.9 | 5.1 ± 1.7 |
| Lateral e' velocity, cm/s | 11.5 ± 4.0 | 6.6 ± 2.2 |

4 All data reported as mean ± standard deviation. All other values represent total number 5 (percent) of affected individuals. Abbreviations: E/e' = E to septal e' velocity ratio; LA = left 6 atrial; LV = left ventricular; TR = Tricuspid regurgitation.

The UK biobank population for prognostic validation consisted of 27,239 subjects, mean age 66±8 years, 50% male. The mean follow-up duration was 1.9 [0.7-4.4] years with 966 cardiovascular events occurring over this period. Of these subjects, 23,051 (85%) had no previous cardiac history at the time of their inclusion. The baseline characteristics of the UK biobank cohort are presented in Table 2.

**Table 2.** UK biobank characteristics.

| **Characteristics** | **UK Biobank Population, all subjects** | **Group with no prior history of cardiovascular disease** | **Group with prior history of cardiovascular disease** |
| --- | --- | --- | --- |
| **n (%)** | 27,239 (100) | 23,051 (85) | 4,188 (15) |
| **Age, years** | 66 ± 8 | 66 ± 8 | 69 ± 7 |
| **Height, cm** | 170 ± 9 | 170 ± 9 | 171 ± 9 |
| **Weight, kg** | 76 ± 15 | 75 ± 15 | 78 ± 15 |
| **Body Surface Area, m<sup>2</sup>** | 1.89 ± 0.22 | 1.88 ± 0.22 | 1.92 ± 0.22 |
| **Male sex, n (%)** | 13,528 (50) | 11,015 (48) | 2,513 (60) |
| **Family History of Cardiovascular Disease, n (%)** | 11,760 (43) | 9,755 (42) | 2,005 (48) |
| **Body Mass Index, kg/m<sup>2</sup>** | 26.5 ± 4.5 | 26.4 ± 4.4 | 27.2 ± 4.6 |
| **Physically Inactive, n (%)** | 2,994 (11.0) | 2,458 (10.7) | 536 (12.8) |
| **Diabetes, n (%)** | 1,667 (6.1) | 1,188 (5.2) | 479 (11.4) |
| **Hypertension, n (%)** | 8,030 (29.5) | 5,833 (25.3) | 2,197 (52.5) |
| **Hyperlipidaemia, n (%)** | 5,548 (20.4) | 3,650 (15.8) | 1,898 (45.3) |
| **Medications, n (%)** | - | - | - |
| **Lipid-lowering** | 4,590 (16.9) | 3,109 (13.5) | 1,481 (35.4) |
| **Antihypertensives** | 4,120 (15.1) | 2,871 (12.5) | 1,249 (29.8) |
| **Insulin** | 134 (0.5) | 94 (0.4) | 40 (1.0) |
| **Any of the three** | 6,120 (22.5) | 4,383 (19.0) | 1,737 (41.5) |
| **Smoking, n (%)** |  |  |  |
| **Current** | 857 (3.1) | 713 (3.1) | 144 (3.4) |
| **Previous** | 9,185 (33.7) | 7,575 (32.9) | 1,610 (38.4) |
| **Never** | 17,098 (62.8) | 14,681 (63.7) | 2,417 (57.7) |
| **Any alcohol, n (%)** | - | - | - |
| **Current** | 25,351 (93.1) | 21,531 (93.4) | 3,820 (91.2) |
| **Previous** | 1,025 (3.8) | 818 (3.5) | 207 (4.9) |
| **Never** | 846 (3.1) | 687 (3.0) | 159 (3.8) |

2 All data reported as mean ± standard deviation. All other values represent total number 3 (percent) of affected individuals.

### Advanced-ECG Score

ECG feature selection identified an optimal A-ECG score containing 7 ECG measures. These ECG measures are detailed in Table 3 including descriptive statistics for these ECG measures on a group level between the normal and diastolic dysfunction groups. This 7-measure score provided excellent discrimination in the derivation cohort (AUC 0.89 [0.86-0.92], as well as in the validation cohort (AUC 0.87 [0.86-0.88], see Table 4 for detailed performance markers). Figure 1 shows representative ECGs from two patients with high (71%) and low (25%) A-ECG scores for diastolic dysfunction, respectively. Sensitivity analyses were undertaken by the addition of patients with grade 1 diastolic dysfunction to the validation cohort. The A-ECG score retained its ability to identify grade 2 and 3 diastolic dysfunction from patients with grade 1 diastolic dysfunction and normal diastolic function with a robust albeit attenuated discriminative ability (AUC 0.78 [0.76-0.80]).

**Figure 1.**
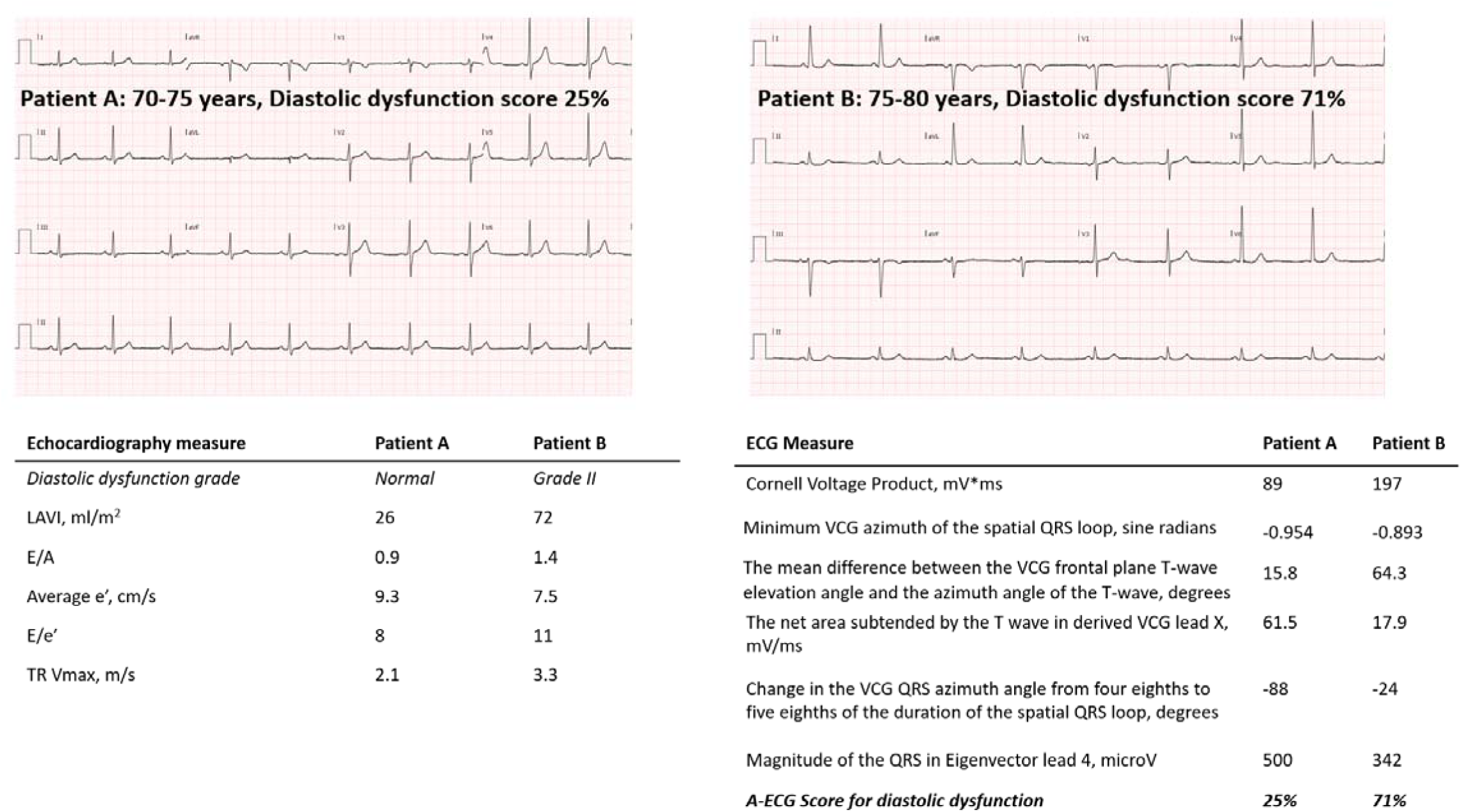
Comparison of A-ECG results from the 12-lead ECGs of two representative patients. Patient A has normal diastolic function with a corresponding low probability of diastolic dysfunction (25%) by A-ECG scoring. Patient B has diagnosed diastolic dysfunction with a high probability of diastolic dysfunction (71%) by A-ECG scoring. Vectorcardiographic (VCG) measures and waveform complexity measures were measured in VCG leads and eigenvector leads, respectively, that were derived from the standard 12-lead ECG.

**Table 3.** A-ECG measures involved in the respective scores. Coefficient estimates derived from elastic net logistic regression, with standardized estimates to allow for approximation of relative contribution to the overall score. Median [IQR] values for each coefficient in the normal diastolic function and diastolic dysfunction cohorts.

| Feature | Coefficient | Standardized Coefficient | Values for normal diastolic function group, n=2649 | Values for grade 2-3 diastolic dysfunction group, n=769 |
| --- | --- | --- | --- | --- |
| Age, years | 0.1186817 | 1.38 | 52 [37–65] | 76 [66–83] |
| Cornell voltage product, mV*ms | 0.0057002 | 0.31 | 110 [77–153] | 141 [96–193] |
| Change in the VCG QRS azimuth angle from four eighths to five eighths of the duration of the spatial QRS loop, degrees | 0.0041626 | 0.13 | -43 [-66 – -24]] | -33 [-55 – -16] |
| The net area subtended by the T wave in derived VCG lead X, mV/ms | -0.0081 | -0.12 | 24.7 [15.5–34.4] | 15.0 [2.5–27.7] |
| The mean difference between the VCG frontal plane T-wave elevation angle and the azimuth angle of the T-wave, degrees | 0.0023645 | 0.07 | 32 [20–50] | 54 [34–86] |
| Minimum VCG azimuth of the spatial QRS loop, sine radians | 0.4874078 | 0.06 | -0.98 [-1 – -0.92] | -0.98 [-1 – -0.92] |
| Magnitude of the QRS in Eigenvector lead 4, microV | 0.0002731 | 0.05 | 391 [297–523] | 442 [329–610] |
| (Intercept) | -8.8852560 | - | - | - |

**Table 4.** Diagnostic Accuracies and Predictive Values of calculated A-ECG Score to identify grade 2+3 diastolic function.

| Performance Marker | Derivation cohort | Validation cohort and sensitivity analysis cohorts |  |
| --- | --- | --- | --- |
|  | Normal +<br>Grade 2-3 DD | Normal +<br>Grade 2-3 DD | Normal +<br>Grade 1-3 DD |
| AUC [95% confidence interval] | 0.89 [0.86-0.92] | 0.87 [0.86-0.88] | 0.78 [0.76-0.80] |
| Sensitivity, % | 73 | 62 | 62 |
| Specificity, % | 89 | 89 | 77 |
| Positive predictive value, % | 80 | 63 | 35 |
| Negative predictive value, % | 85 | 89 | 91 |
| Positive likelihood ratio | 6.9 | 5.8 | 2.7 |
| Inverse negative likelihood ratio | 3.3 | 2.3 | 2.0 |

Abbreviations: DD = diastolic dysfunction, AUC = Area under the receiver operating characteristics curve

### Prognosis in the UK Biobank

Prognostic validation of the score was conducted in a cohort selected from the UK Biobank. Subjects divided into quartiles based on their A-ECG score demonstrated increasing risk of cardiovascular events with increasing score (log-rank 338, p<0.001, Figure 2). When compared to those with scores ≤25%, subjects with a score >75% had a hazard ratio (HR) of 5.7 [4.5-7.2] (p<0.001). When adjusted for age, sex and co-morbidities the HR was 1.9 [1.4-2.6, p<0.001).This elevated risk persisted when patients were divided into subgroups with no prior diagnosis of cardiovascular disease and those with known cardiovascular disease (HR 5.5 [4.0-7.5], p<0.001, and HR 3.3 [2.3-4.5], p<0.001, respectively for diastolic dysfunction scores >75%). A comparison of hazard ratios by quartile of A-ECG score is outlined in Table 5. Survival analysis using spline modelling showed that over the range of the score (0-100%), hazard ratios varied linearly between 1 and 15 (p<0.001, Supplementary Figure 4). After adjusting for age, sex, and cardiovascular risk factors this linear relationship persisted, for hazard ratios ranging between 1 and 4 (p<0.001, Figure 2).

**Figure 2.**
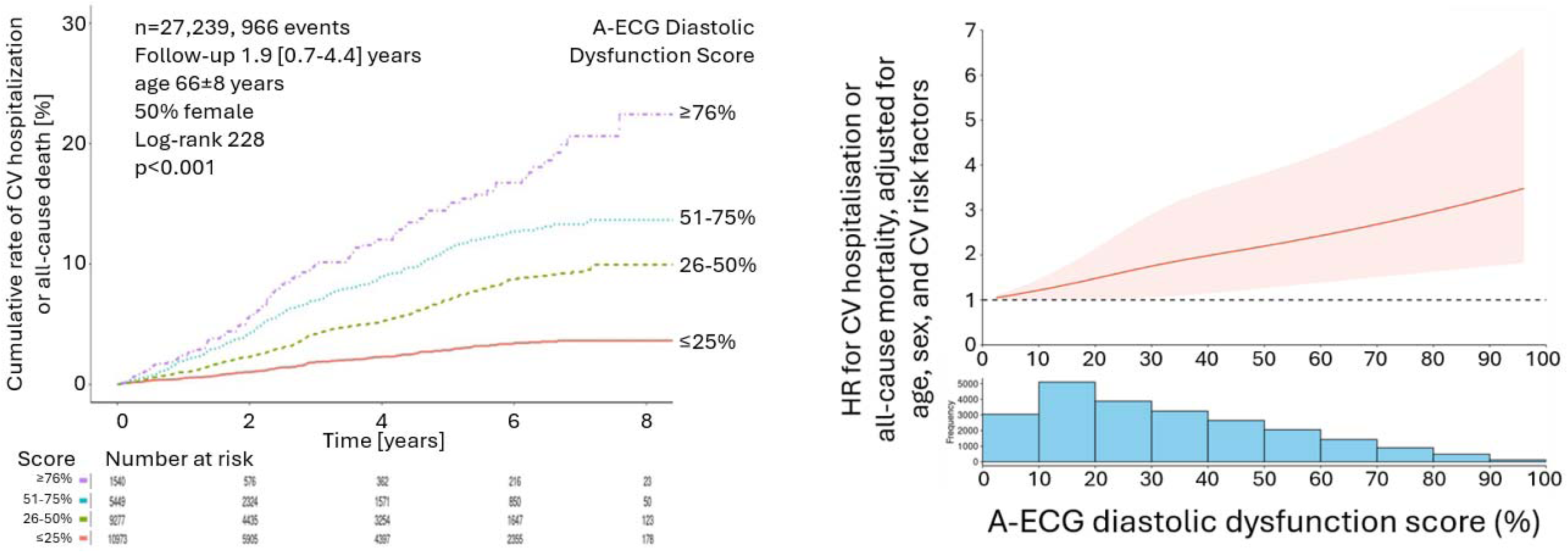
Cumulative event curves per quartile of A-ECG diastolic dysfunction score (left). Restricted cubic spline of adjusted hazard ratio for CV hospitalisation and all-cause mortality (right). Note the ‘dose-response’ relationship of increasing events with increasing A-ECG diastolic dysfunction score, even after adjusting for age, sex, and cardiovascular risk factors.

**Table 5.** Comparison of hazard ratio by A-ECG diastolic dysfunction score quartile.

**Full cohort, n=27,239**
| **A-ECG Diastolic Dysfunction Score, %** | **Hazard ratio [95%CI] vs score <25%** | **p value** |
| --- | --- | --- |
| 25–50 | 2.5 [2.1–3.0] | < 0.001 |
| 50–75 | 4.0 [3.3–4.7] | < 0.001 |
| ≥75 | 5.7 [4.5–7.2] | < 0.001 |

**No prior known cardiovascular disease, n=23,051**
| **A-ECG Diastolic Dysfunction Score, %** | **Hazard ratio [95%CI] vs score <25%** | **p value** |
| --- | --- | --- |
| 25–50 | 2.6 [2.1–3.2] | < 0.001 |
| 50–75 | 4.0 [3.2–5.1] | < 0.001 |
| ≥75 | 5.5 [4.0–7.5] | < 0.001 |

**Prior known cardiovascular disease, n=4,188**
| **A-ECG Diastolic Dysfunction Score, %** | **Hazard ratio [95%CI] vs score <25%** | **p value** |
| --- | --- | --- |
| 25–50 | 1.8 [1.4–2.4] | < 0.001 |
| 50–75 | 2.4 [1.8–3.2] | < 0.001 |
| ≥75 | 3.3 [2.3–4.7] | < 0.001 |

## Discussion

The major finding of this study is that an explainable 7-measure A-ECG score derived from a standard 12-lead ECG can identify diastolic dysfunction with excellent diagnostic and strong prognostic performance (Central Illustration). The score’s reported likelihood ratios demonstrate the utility of A-ECG analysis on an individual level independent of the prevalence of diastolic dysfunction in the population being studied. For example, the score’s positive likelihood ratio of 5.8 suggests that following a positive A-ECG result, a patient is 5.8 times more likely to be diagnosed with diastolic dysfunction on TTE. Furthermore, the score’s prognostic performance in the UK biobank population illustrates that in addition to diagnostic performance for identifying diastolic dysfunction at a cutoff of 50% probability, the score as a continuous variable ranging from 0-100% probability associates linearly with cardiovascular events and all-cause mortality even after adjusting for age, sex, and cardiovascular risk factors. This further strengthens the utility of A-ECG as a low-cost screening tool for identifying diastolic dysfunction in at-risk populations potentially amenable to targeted prevention before development of HFpEF.

### Validity and feasibility of A-ECG analyses

The introduction of the ECG in 1902 provided, for the first time, objective information about the electrical activity and function of the heart that could subsequently be used to diagnose a number of cardiac pathologies such as atrial fibrillation and myocardial infarction^25,26, 27^. However, it was only in the 1960s that a multivariable approach to the conventional ECG analysis was applied to improve diagnostic detection of diseases such as LV hypertrophy and prior myocardial infarction^28, 29^. A-ECG analysis is an extension of this arc of development, and is able to extract diagnostic and prognostically valuable information from a standard resting 12-lead ECG for the detection of a large range of cardiac pathologies^11^. It has been shown to improve diagnostic accuracy when compared to conventional ECG in coronary artery disease, cardiomyopathies, LV hypertrophy and LV systolic dysfunction^11, 14, 15^.

To date, there is no widely accepted ECG-based method for detecting diastolic dysfunction. A machine learning model using clinical and ECG variables has shown potential to detect diastolic dysfunction with an AUC of 0.80 in an external test cohort^7^. However, that model relied on inclusion of a range of information including clinical co-morbidities such as hypertension and dyslipidaemia that may limit its applicability as a screening tool when such information is not readily available. Signal-processed ECG analysis has also demonstrated utility in in detecting reduced mitral annular velocities, however data evaluating that method have been limited to feasibility assessments with small patient cohorts^30^. A single-lead ECG recorded for 3 minutes using specialised dedicated hardware and a sampling rate far exceeding routine clinical ECG has also been able to achieve high diagnostic accuracy for detecting diastolic dysfunction^8^. However, a major barrier for widespread use of that method as a routine screening tool in large populations is the need for dedicated hardware. Artificial intelligence (AI) derived models have also shown promise for both diagnosing diastolic dysfunction and assessing prognostic performance^9, 31^. However, a key limitation of AI-derived models is their ‘black box’ nature, providing limited insight into the ECG features that underpin their diagnostic performance, which may compromise clinical adoption. By comparison, A-ECG analysis can be performed on a digital recording from a standard, 10-second, resting 12-lead ECG recorded using existing clinical standard-fidelity ECG devices^11^. In contrast to AI models, A-ECG is advantageous as features contributing to the score are transparently known and readily explainable, which may not only improve generalizability but also help drive both clinical acceptance as well as an understanding of the underlying mechanisms of diastolic dysfunction. Notably A-ECG diagnostic techniques are already used and reimbursed clinically in New Zealand.

### Components of the score

The Cornell voltage product was the single strongest performing A-ECG feature in the diastolic dysfunction score. This is consistent with prior research which suggests that an increased Cornell voltage product could be used to help diagnose diastolic dysfunction and was a marker of poor prognosis^32^. The score also includes different vectorcardiographic measures of the spatial representation of the QRS complex as a loop in three-dimensional space. Indeed, such measures have been shown to have diagnostic and prognostic power in cardiovascular disease states including other heart failure phenotypes^33, 34^.

Two of the six A-ECG features included in the score examined the T wave morphology. Differences in T wave characteristics and the QT duration have also been associated with the presence of diastolic dysfunction. It is believed that the progressive myocardial structural changes seen in diastolic dysfunction impact calcium cycling during ventricular relaxation which can have subtle changes detectable in the 12-lead ECG^35, 36^.

As such, the current study shows that the use of an A-ECG score for detecting diastolic dysfunction achieves excellent diagnostic performance with methods that are economically and logistically feasible for widespread clinical use. In addition to its diagnostic utility, our validation demonstrates that an elevated A-ECG diastolic score carries prognostic information along a continuum of probability (0-100%), independent of risk factors for cardiovascular disease in a large out-patient population with low prevalence of prior known cardiovascular disease. Indeed, the score had the strongest association with prognosis among those with no prior known cardiovascular disease. Furthermore, A-ECG analysis can be performed automatically, rapidly, and inexpensively using a standard resting 12-lead ECG while at the same time also diagnosing or screening for other pathologies^37^. Moreover, it is likely that many patients at risk of having or developing diastolic dysfunction would have pre-existing resting 12-lead ECGs from the past for convenient retrospective analysis.

### Practical applications of A-ECG scores

In the future, A-ECG may have application in a range of different clinical settings, in particular in primary care where this novel information may allow for more accurate risk stratification for cardiac disease and assist in identifying only those patients who might most benefit from further cardiac imaging. In addition, the low skill and cost required to perform this analysis could provide substantial value in population-based settings wherein relatively technically complex and expensive methods such as echocardiography might not be feasible. Another beneficial aspect of A-ECG is that it can simultaneously provide probability scores for a wide range of cardiac disease states ^37^. Upon knowing the results of transparently explainable numeric probability scores available at the time of consultation, physicians may be better equipped to engage with patients regarding preventative therapy and lifestyle changes to reduce the risk of developing cardiovascular disease. Diastolic dysfunction is under-diagnosed and a precursor to clinically overt HFpEF, the leading cause of heart failure^38^. As such, a convenient and low-resource screening tool for the diagnosis of diastolic dysfunction would be particularly valuable to aid early identification of individuals who might benefit from further cardiac assessment and risk factor modification to prevent or delay the development of HFpEF. Additional, fully prospective follow-on studies using the proposed optimal A-ECG score are therefore justified.

### Limitations

The database used to train and validate this model had limited demographic information on patients, including a lack of information on the prevalence of cardiovascular risk factors such as diabetes and hypertension. Given that the control group was recruited from patients referred for outpatient cardiac assessment, it is also likely that this population contains a higher prevalence of cardiac co-morbidities than what would be expected from asymptomatic, healthy controls. It is also likely that even age-matched healthy controls would display an increased probability of diastolic dysfunction due to normal healthy ageing. Despite these potential shortcomings, the score demonstrated both robust diagnostic performance and prognostic performance in independent datasets. The diagnostic performance validation was performed in an external cohort selected from another institution in a different country, and among patients with broad inclusion criteria, supporting its utility in other populations. The prognostic performance in the UK Biobank underscores utility in an outpatient population that was largely without prior known cardiovascular disease, highlighting its utility in such populations moving forward.

The strength of any predictive tool is dependent on the population tested. The utility of the score might also be limited in patients underrepresented in our study cohort, in particular patients younger than 50 years of age. However, it is worth noting that younger patients are less likely to be a target population for screening for diastolic dysfunction. Furthermore, only patients in sinus rhythm and without significant conduction disease were included in the study. Patients with atrial arrhythmia and/or ventricular conduction disease, both prevalent in patients with diastolic dysfunction and heart failure, would therefore not be screened using the proposed A-ECG score. It would therefore be beneficial to develop additional, dedicated scores for patients with atrial fibrillation or advanced conduction disease in the future. Whilst this study did not include any patients in atrial fibrillation or flutter, our 7-measure score did not use any P-wave derived measures and demonstrated excellent diagnostic accuracy.

Whether this score might therefore have diagnostic utility in patients with atrial arrhythmias remains to be studied. Age is a feature of the score, and diastolic dysfunction prevalence is known to increase with age^5^. By comparison, sex did not make it into the score. Despite age being included in the score, the score retained a robust association with prognosis in the UK Biobank population when adjusted for age, sex, and cardiovascular risk factors. This indicates that age is not the sole driver of the multivariable score, and the ECG features provide unique information.

## Conclusions

An inherently explainable and transparent score derived from A-ECG analysis of the standard resting 12-lead ECG is able to detect diastolic dysfunction with high diagnostic accuracy and clinically meaningful prognostic performance along a continuum of probability. A-ECG shows potential as a screening tool for diastolic dysfunction, to potentially optimal use of echocardiography while facilitating early management of cardiovascular risk factors to help mitigate progression to HFpEF.

## Data Availability

The data underlying this article can be made available upon reasonable request to the corresponding author.

---

**Central illustration:**
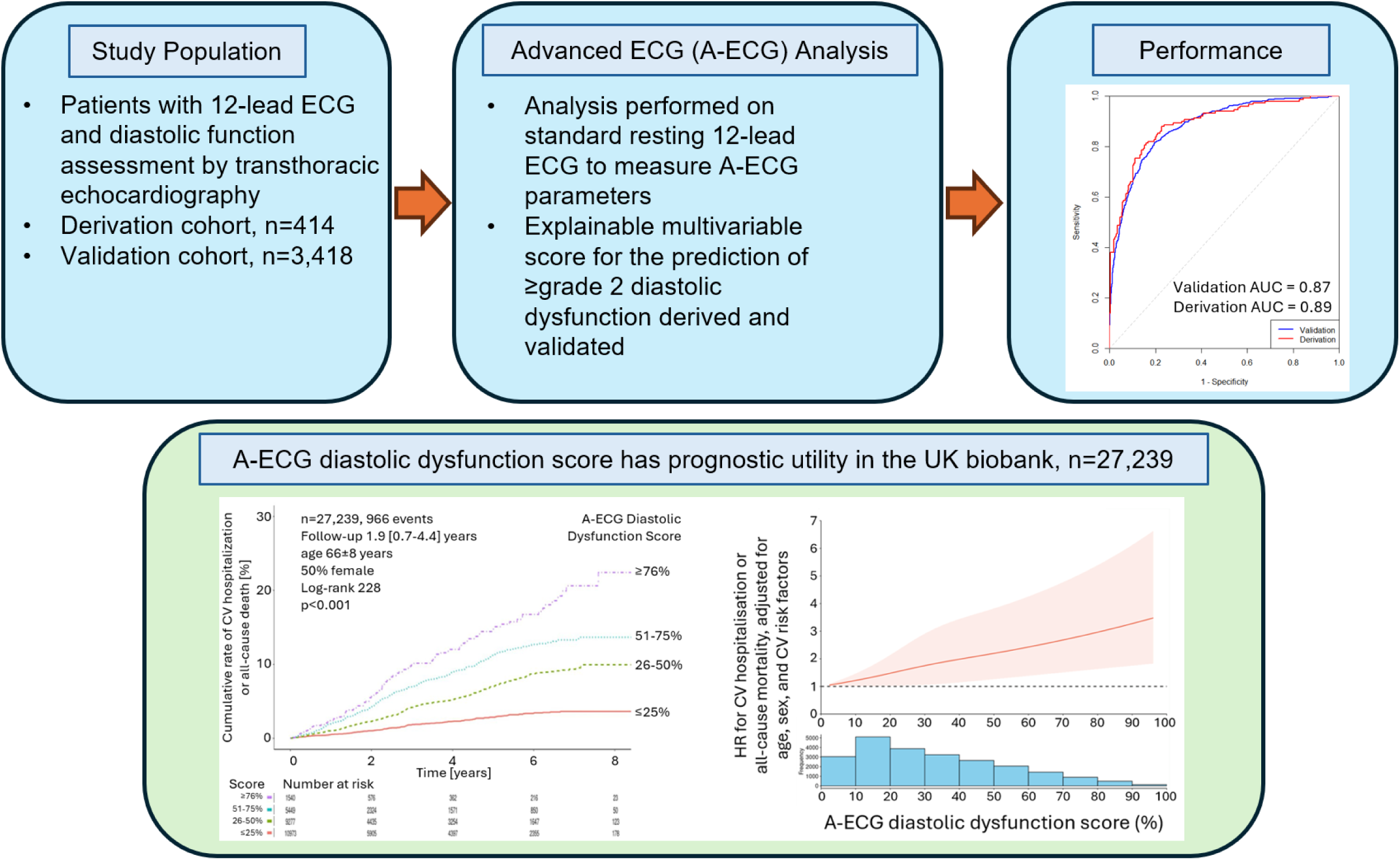
An Explainable Advanced Electrocardiography Score for Diastolic Dysfunction. Summary of study design and main findings of the article.

