## Supplementary material for "An Explainable Advanced Electrocardiography Score for Diastolic Dysfunction - Derivation, Validation and Prognostic Performance"

**Supplementary Table 1: ICD-10 diagnoses and OPCS-4.10 codes used to define cardiovascular events**
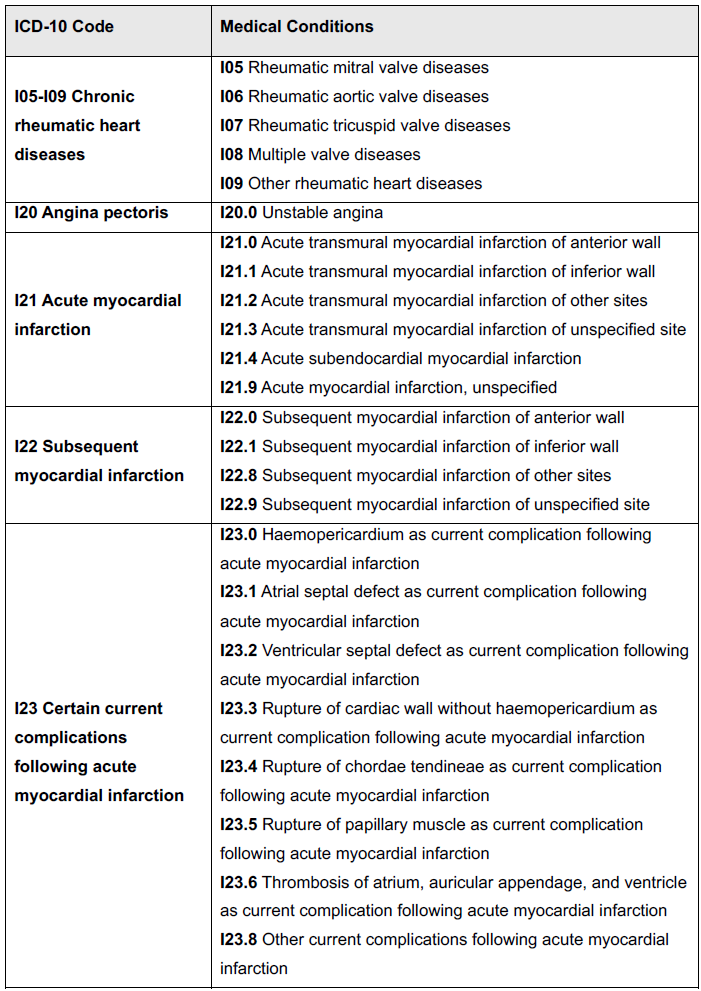

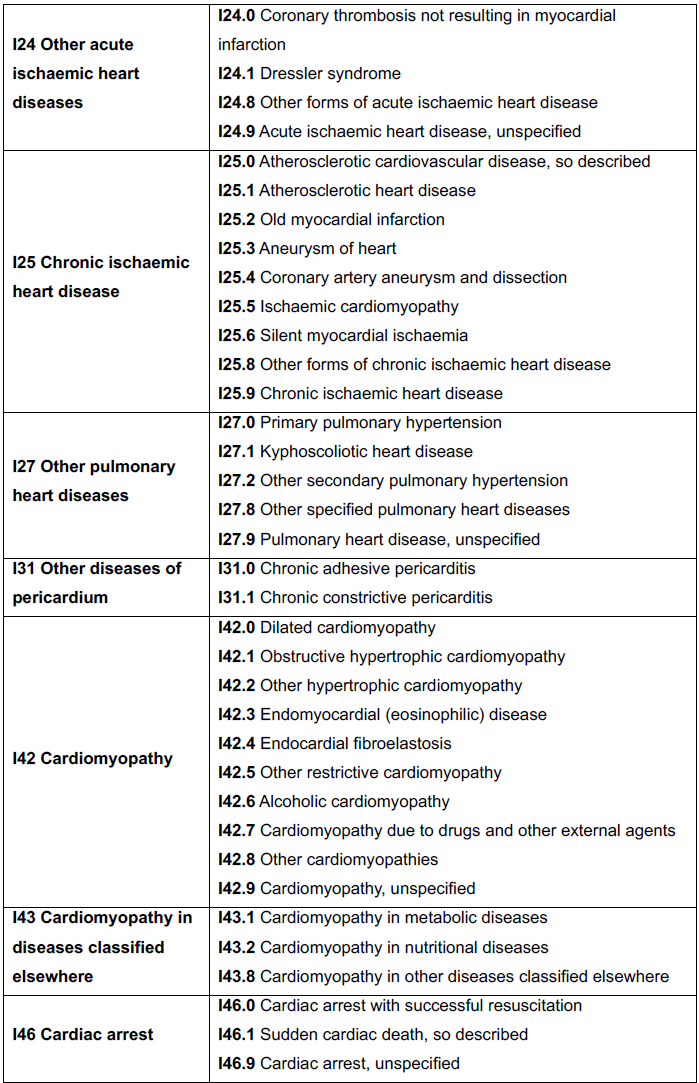

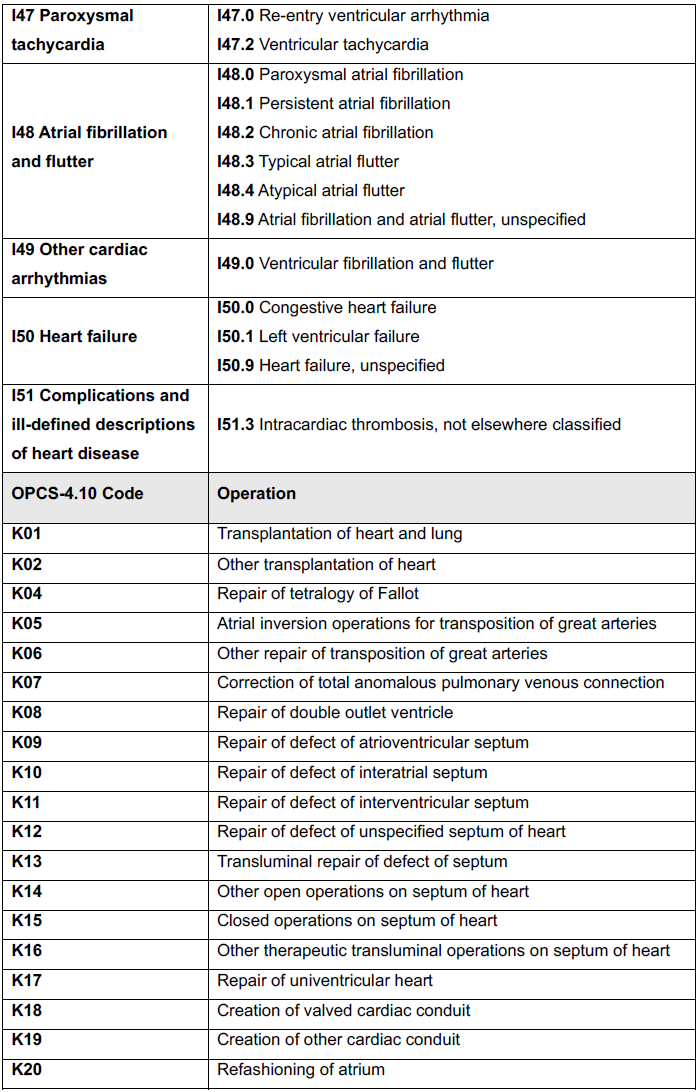

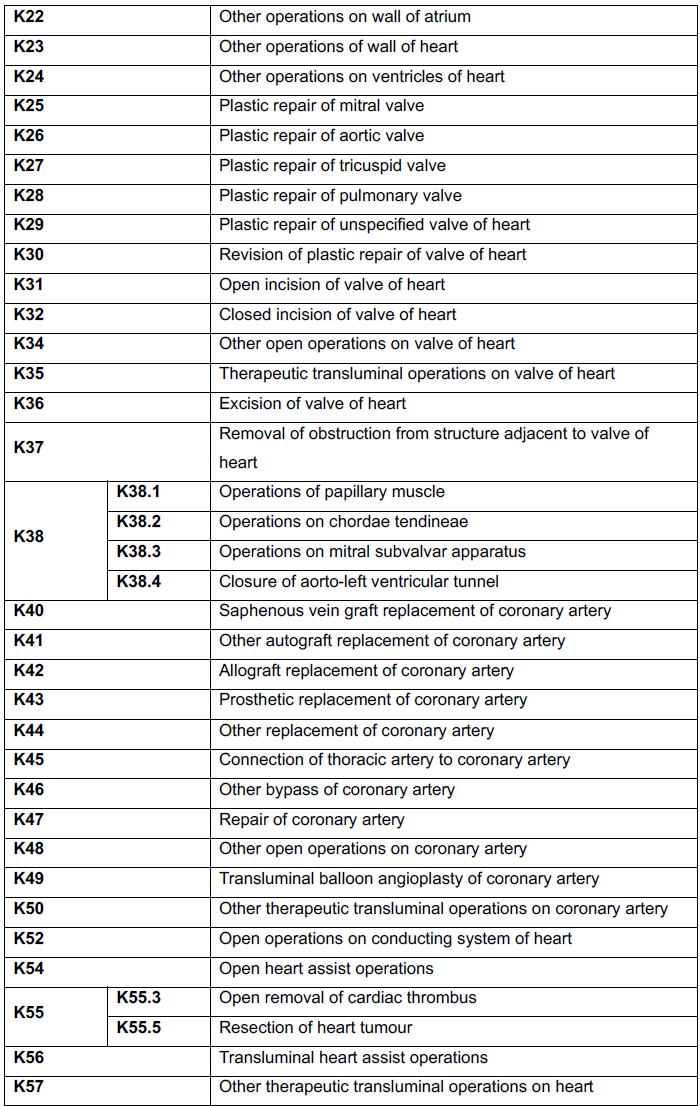

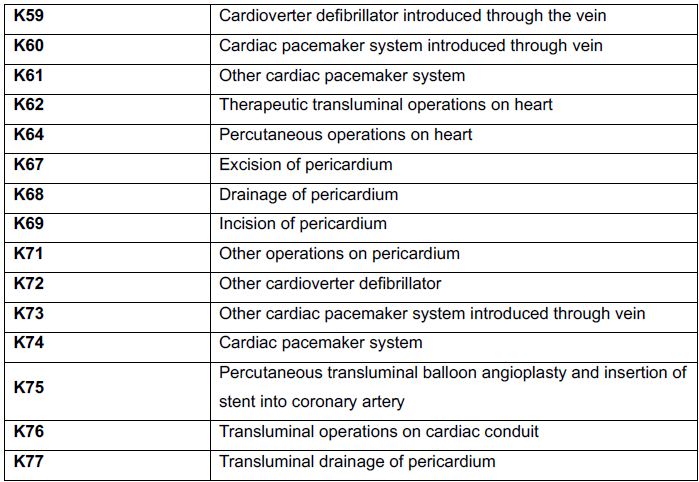

Table 1 outlines the ICD-10 diagnosis codes as provided by World Health organisation and the OPCS-4.10 operation codes as provided by NHS England that were used to define cardiovascular events.

**Supplementary Table 2. List of ICD-10 diagnosis used to define the presence of pre-existing cardiovascular disease**

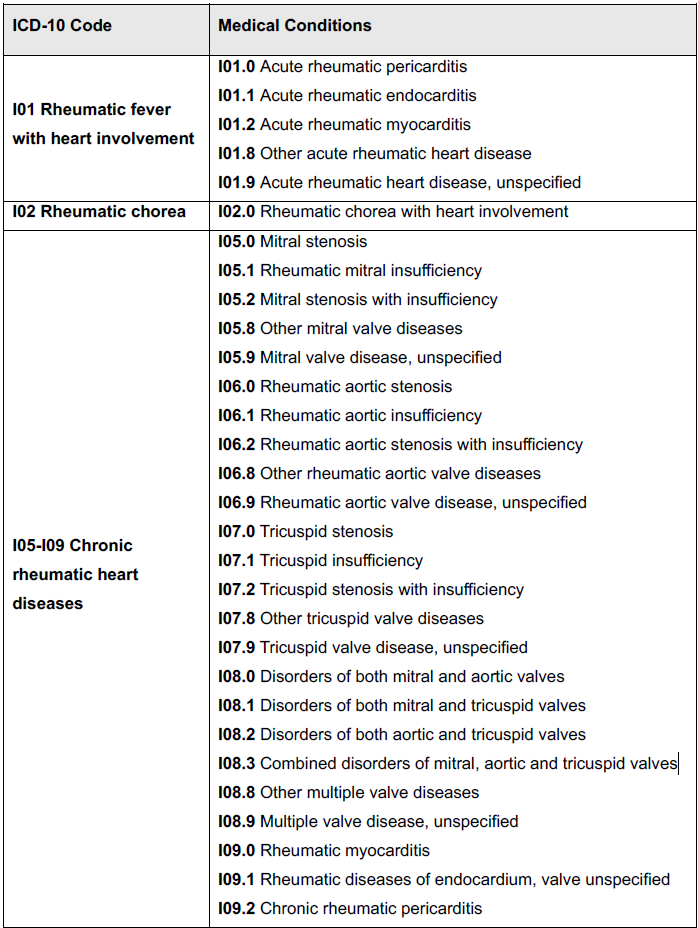

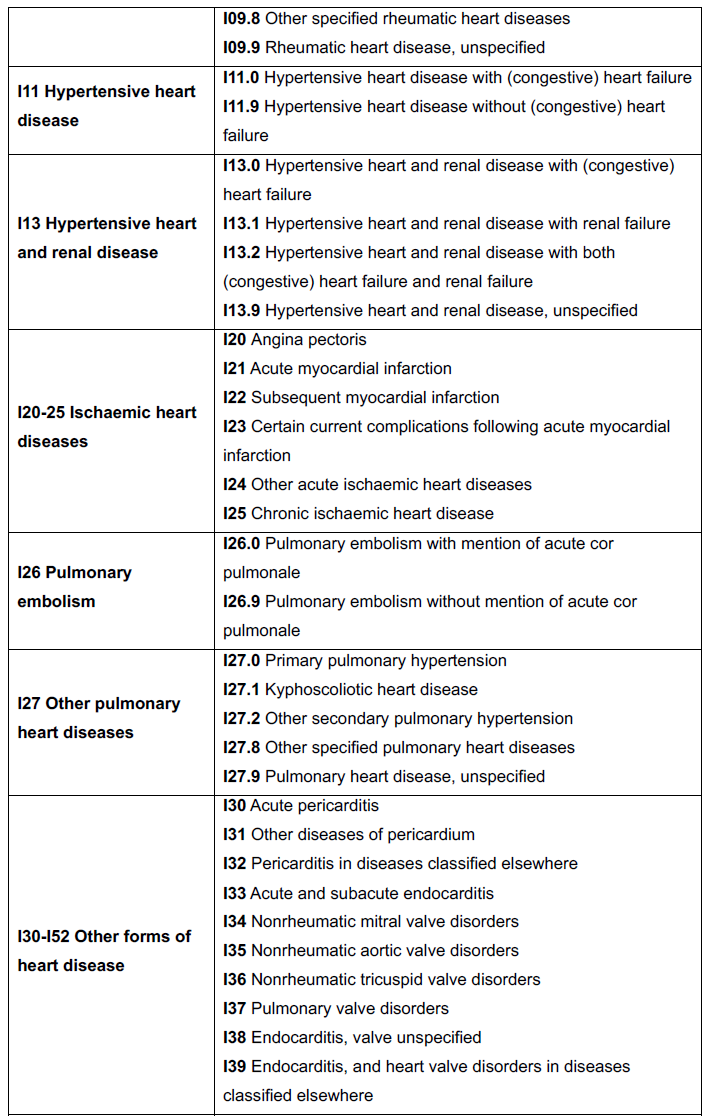

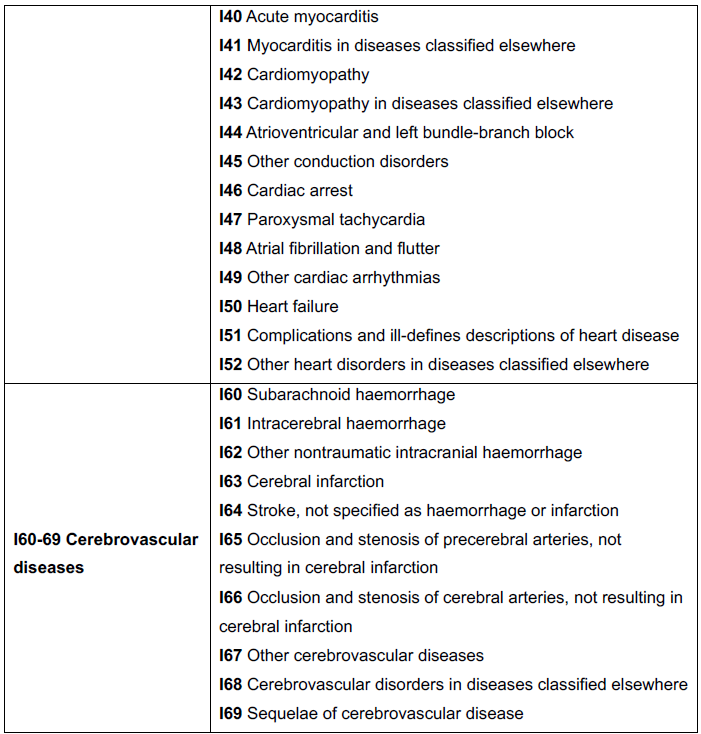

**Supplementary Table 3. Derivation Cohort Patient characteristics**

| **Variable** | **Normal diastolic function** | **Diastolic dysfunction** |
| --- | --- | --- |
| Number of patients, n (%) | 264 (64) | 150 (36) |
| Male sex, n (%) | 119 (45) | 53 (35) |
| Age, years | 67.9 [60.6–75.2] | 83.2 [78.1–89.2] |
| Body surface area, m^2^ | 1.9 ± 0.2 | 1.8 ± 0.2 |
| Echocardiographic data | | |
| LA volume index, ml/m^2^ | 29.8 ± 8.0 | 45.2 ± 10.5 |
| LV septal wall thickness, mm | 9.4 ± 1.8 | 11.3 ± 2.1 |
| LV posterior wall thickness, mm | 9.1 ± 1.0 | 10.0 ± 2.2 |
| E/A ratio | 1.1 ± 0.4 | 1.1 ± 0.5 |
| Average E/e’ ratio | 8.3 ± 2.3 | 14.9 ± 4.4 |
| TR velocity, m/s | 2.3 ± 0.3 | 2.8 ± 0.5 |
| Peak E-Wave velocity, cm/s | 71 ± 17 | 88 ± 21 |
| Average e’ velocity, cm/s | 9.1 ± 2.6 | 6.3 ± 2.0 |
| Septal e’ velocity, cm/s | 10.6 ± 3.2 | 7.1 ± 2.5 |
| Lateral e’ velocity, cm/s | 7.6 ± 2.2 | 5.7 ± 1.8 |

Data reported as mean ± standard deviation or Median [IQR] as appropriate. All other values represent total number (percent) of affected individuals. Abbreviations: E/e’ = E to septal e’ velocity ratio; LA = left atrial; LV = left ventricular; TR = Tricuspid regurgitation

**Supplementary Table 4. Diagnostic validation cohort characteristics based on diastolic dysfunction grade**

| **Variable** | Grade 1 | Grade 2 | Grade 3 |
| --- | --- | --- | --- |
| Number of patients, n (%) | 1198 (61) | 690 (35) | 79 (4) |
| Male sex, n (%) | 447 (37) | 230 (33) | 50 (63) |
| Age, years | 72.2 ± 10.6 | 74.3 ± 13.0 | 71.1 ± 14.7 |
| Body surface area, m^2^ | 1.8 ± 0.2 | 1.8 ± 0.3 | 1.9 ± 0.3 |
| **Echocardiographic data** | | | |
| Ejection fraction (%) | 60.2 ± 8.5 | 59.0 ± 11.4 | 44.7 ± 15.7 |
| LA volume index, ml/m^2^ | 34.3 ± 9.9 | 46.8 ± 12.4 | 54.6 ± 17.1 |
| LV septal wall thickness, mm | 11 ± 3 | 11 ± 3 | 11 ± 2 |
| LV posterior wall thickness, mm | 9 ± 2 | 9 ± 3 | 10 ± 2 |
| E/A ratio | 0.6 ± 0.1 | 1.0 ± 0.3 | 3.5 ± 1.9 |
| Average E/e’ ratio | 9.3 ± 2.5 | 15.1 ± 6.2 | 15.6 ± 8.3 |
| TR velocity, m/s | 2.4 ± 0.2 | 2.9 ± 0.4 | 3.2 ± 0.5 |
| Peak E-Wave velocity, cm/s | 52.9 ± 12.5 | 90.8 ± 24.5 | 105.0 ± 25.2 |
| Septal e’ velocity, cm/s | 5.0 ± 1.2 | 5.1 ± 1.7 | 5.4 ± 1.7 |
| Lateral e’ velocity, cm/s | 6.3 ± 1.7 | 6.5 ± 2.2 | 7.5 ± 2.5 |

**Supplementary Figure 1. Flowchart describing derivation cohort inclusion and exclusion criteria**

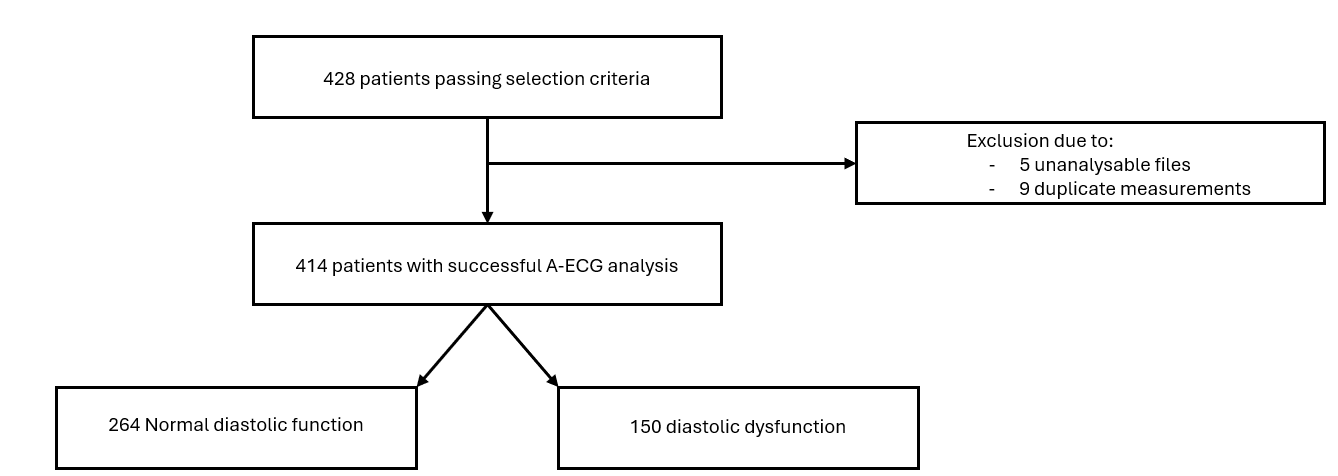

The selection criteria included patients referred for assessment at a large academic outpatient cardiology practice. The inclusion criteria included age >50, sinus rhythm, LVEF >55% and a recorded 12-lead ECG within 1 month of TTE. Patients were excluded if they had moderate or greater valvular disease, LVEF <55%, hypertrophic cardiomyopathy, valvular replacement, coronary bypass surgery, or any other cardiac procedures that required entering the pericardium. ECG abnormalities that would limit A-ECG analysis such as atrial arrhythmias, predominant ventricular ectopic beats and QRS duration >120ms were also excluded. Abbreviations: LVEF = Left ventricular ejection fraction; TTE = Transthoracic echocardiography.

**Supplementary Figure 2. Flowchart describing diagnostic validation cohort inclusion and exclusion criteria**

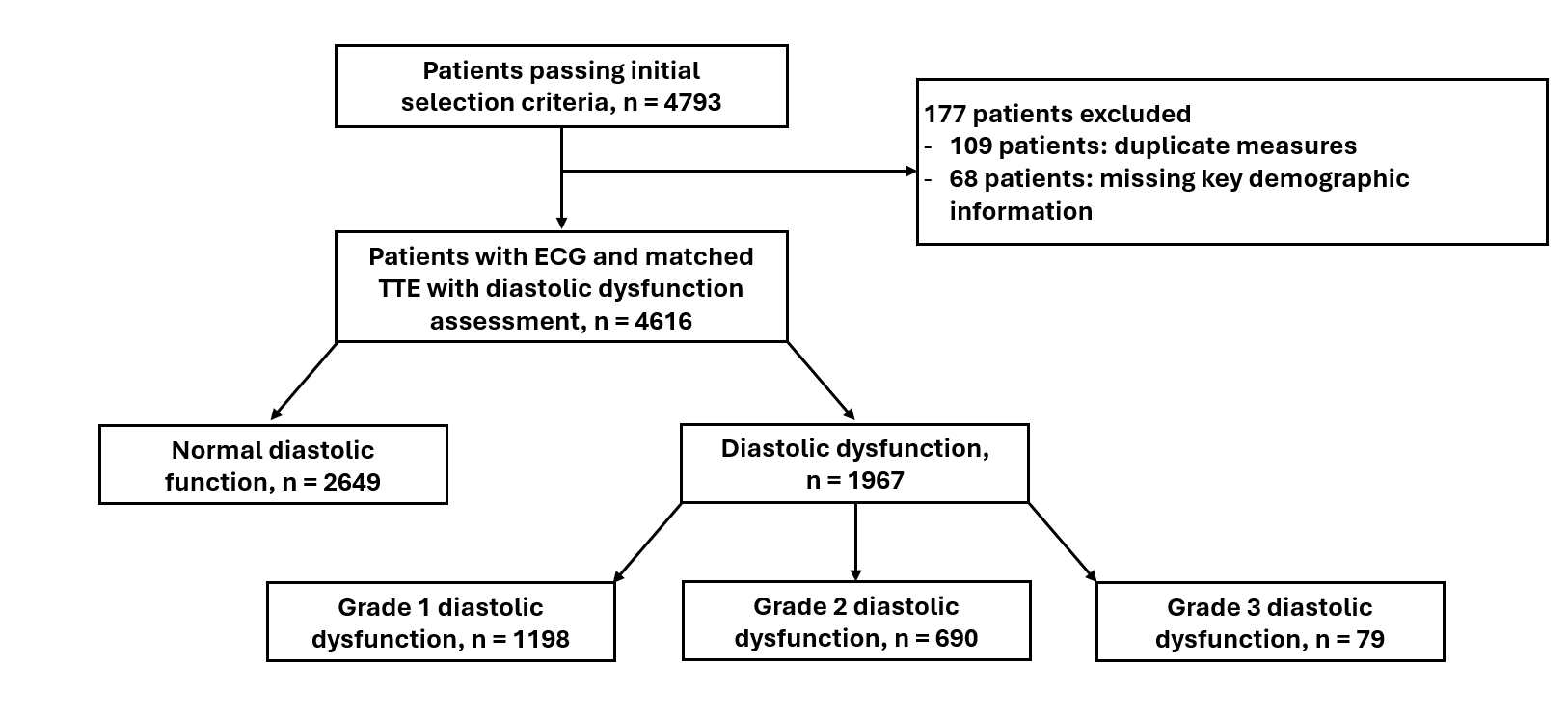

The selection criteria included patients with a recorded 12 lead ECG and a transthoracic echocardiogram with diastolic function assessment.

**Supplementary Figure 3 – Flowchart describing inclusion and exclusion criteria of the UK biobank prognostic validation cohort**

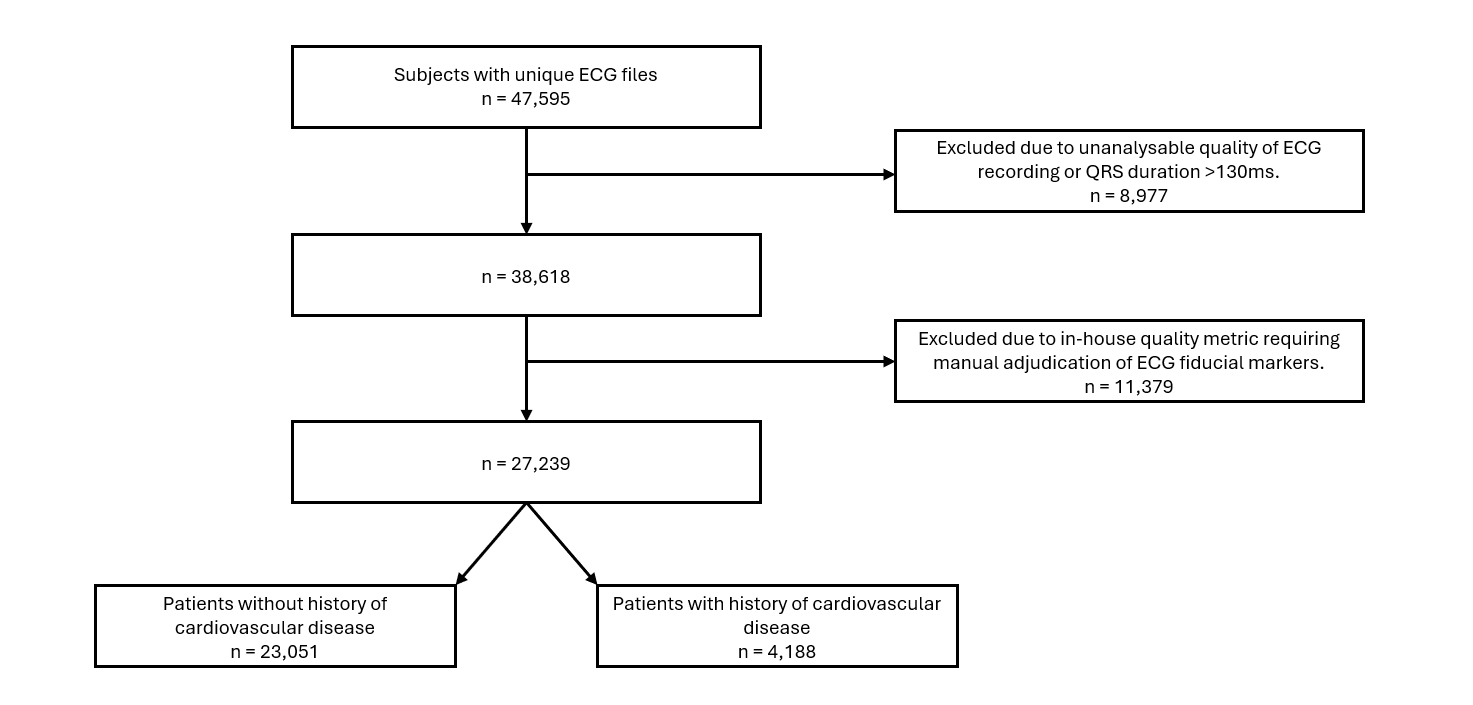

**Supplementary Figure 4 – Restricted cubic spline of unadjusted hazard ration for CV hospitalisation or all cause mortality by A-ECG diastolic dysfunction score**

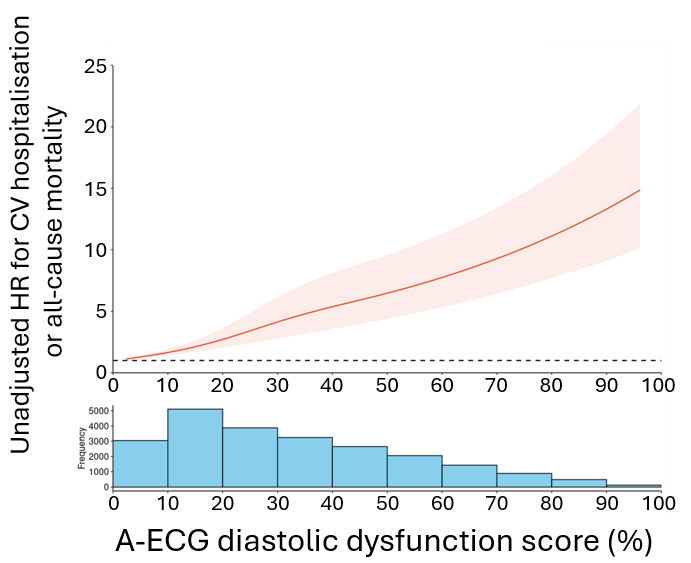
